# Insights on the Implementation Strategies for a Potential New Tuberculosis Vaccine in South Africa

**DOI:** 10.1101/2025.08.01.25332725

**Authors:** Katherine A. Thomas, Choice Okoro, Thobani Ntshiqa, Verena Damovsky, Deanne Goldberg, Omphile Ramokhoase, Rebecca A Clark, Waasila Jassat, Gavin J Churchyard, Fareed Abdullah, Mmamapudi Kubjane, Janet Seeley, Salome Charalambous, Richard G White

## Abstract

**Background:** Several tuberculosis (TB) vaccines intended for adults and adolescents are in late-stage clinical trials, but there is limited research into how a new TB vaccine would be introduced. South Africa is at the forefront of TB vaccine research, with involvement in the clinical trials of leading candidates, in this study; we sought to understand what the interest, priorities and roll out approach of a potential new TB vaccine in South Africa would be.

**Methods:** We conducted semi-structured interviews with stakeholders with different expertise to understand their approach to the implementation of a new TB vaccine. A total of 26 stakeholders were interviewed during April and May 2025. Deductive analysis was used to develop a coding framework for thematic content analysis of the data.

**Results:** Stakeholders supported introducing a new TB vaccine in South Africa due to the high disease burden, but feasibility of the roll out was said to depend on vaccine price, cost-effectiveness, efficacy, regulatory approval, and implementation logistics. Efficacy below 50% could raise concerns unless supported by robust modelling demonstrating significant public health impact and cost-effectiveness. Stakeholders preferred a cheaper vaccine course to maximise availability and enable a broad population-based introduction. Priority populations for vaccination included adolescents and adults in the case of a broad population approach, and high-risk groups such as, people living with HIV, healthcare workers, TB household contacts, miners, prisoners and people living with diabetes. Whilst a national broad population-based approach was preferred, constraints may result in a phased rollout in high-burden areas and/or targeted vaccination to high-risk groups first. Strong advocacy, detailed cost effectiveness and cost benefit assessments, regulatory alignment, and integrated service delivery were seen as essential to successful implementation.

**Conclusions:** With the introduction of a new TB vaccine potentially as early as 2030, this study provides valuable insights, particularly regarding target populations, that can be utilised for the implementation of a new TB vaccine in South Africa to ensure its introduction is successful. Further work is required on vaccine acceptability and TB infection prevalence and modelling to estimate cost effectiveness and budget impact.

## Introduction

In 2023, tuberculosis (TB) was the leading cause of death globally by a single infectious agent. Over 10 million people continue to fall ill with TB every year, with this number on the rise [1]. South Africa has a particularly high burden of TB, with an incidence rate of 427 per 100 000 of the population, and around 5% of cases expected to be multidrug resistant (MDR-TB) or rifampicin resistant (RR-TB) [1]. Not only does South Africa have a high burden of TB, but it also carries a substantial burden of human immunodeficiency virus (HIV) at 17.8% of all 15-49 year olds in 2022, contributing to its classification by the World Health Organization (WHO) as one of the countries with the highest burden of HIV-associated TB [2, 3]. Progress toward global TB elimination targets have stalled in recent years, in part due to the COVID-19 pandemic’s disruption of TB services, which was particularly pronounced in high-burden settings like South Africa [1, 4].

The burden of TB is predominantly concentrated among adolescents and adults, who account for the majority of incident TB cases and deaths [5]. Modelling studies have demonstrated that successful vaccination of these age groups, rather than infants, would result in a significantly quicker and greater public health impact [6, 7]. Despite this, the only currently licensed TB vaccine is the Bacillus Calmette-Guérin (BCG) vaccine which is delivered neonatally and protects against miliary and meningeal TB among infants and young children [8]. A recent BCG revaccination trial conducted in South Africa demonstrated that revaccination of BCG in adolescent populations does not provide protection from sustained *Mycobacterium Tuberculosis (Mtb)* infection [9]. In addition, the contraindication of the BCG vaccine among adults living with HIV is a clear potential limitation for use in South Africa due to its high HIV burden, collectively demonstrating the urgent need for novel TB vaccines using different technologies and specifically targeting adolescent and adult populations.

Several TB vaccine candidates are in late-stage clinical trials to estimate both the safety and efficacy among adolescents and adults, candidates include M72/AS01E, MTBVAC and VPM1002, all of which are currently being trailed in South Africa [10, 11]. Among the most advanced is the M72/AS01E adjuvanted recombinant protein vaccine, which previously demonstrated 49.7% (95% CI 2.1 to 74.2) efficacy in preventing pulmonary TB among IGRA+ adults within a Phase IIb trial conducted in Kenya, Zambia and South Africa [12]. With data from a Phase III clinical trial anticipated to be available in 2028, it is plausible that a novel TB vaccine for adolescents and adults could be licensed and available as early as 2030.

As adolescents and adults are not typically the target population for vaccinations, there is limited evidence on how vaccination programmes can be effectively rolled out among this population [5, 13]. New guidance has been produced by WHO on the evidence required for successful introduction of vaccine to adolescents and adults, including the ‘Evidence Considerations for TB Vaccine Policy (ECVP)’ [14] and the ‘WHO Global Framework to Prepare for Country Introduction of New TB Vaccines for Adolescents and Adults’ [15]. Whilst Pelzer et al (2022) [13] provided insights into the implementation strategies for a new TB vaccine in South Africa, their interviews were conducted just as the COVID-19 pandemic began, and as such this research lacks lessons learnt during the COVID-19 vaccination roll out which began in February 2021 [16]. A scoping review of TB vaccination implementation research published in 2024 highlighted the scarcity of research into the implementation strategies emphasizing the need for further research to ensure a rapid, successful and equitable vaccine implementation [17].

In this study we aimed to understand implementation priorities of a novel TB vaccine for adolescents and adults among key stakeholders in South Africa. We conducted interviews with a range of stakeholders to understand the key enablers and barriers of a new TB vaccine roll out, priority implementation strategies and key target populations. These findings could inform new TB vaccine introduction in South Africa, including the development of policy.

## Methods

### Study Design

This research is part of a broader multi-country project aiming to create evidence to support national and global TB decision-makers in reducing the burden of TB. For this research we conducted in-depth interviews among key stakeholders in South Africa. The aim was to evaluate how a new TB vaccine that prevents disease would be introduced within South Africa and to understand the likely target populations of the vaccine and implementation strategies.

Semi-structured interviews were conducted during April and May 2025 and covered a range of themes including country interest in a new TB vaccine, introduction timelines, target populations, regulatory considerations, financing, and lessons from previous health programmes. The interview guide (supplementary file 1) was adapted for each participant depending on their area of expertise.

### Study Population and Setting

South Africa was selected for this study due to its high TB burden and likelihood of early vaccine adoption given its participation in late-stage TB vaccine trials [1]. Using purposive sampling, based on participants from a range of expertise, we recruited 26 participants working in a range of sectors relevant to decision making for potential TB vaccine introduction. Within the National Department of Health (NDoH), we had representation from the TB directorate, Expanded Programme on Immunization (EPI) and Primary Health Care (PHC).

Participants were identified through professional networks and the selection aimed to ensure representation across different areas of TB, vaccine, regulatory and advocacy expertise. Participants were invited by email between 10/04/2025 and 21/05/2025. They were provided with a summary of the study, background on the TB vaccine pipeline, and logistical information. The table of participant expertise is summarised in the Supplementary Table 1.

### Data Collection

Interviews were conducted by two trained researchers with expertise in qualitative research and TB vaccines. We conducted a total of 15 interviews, 10 of which were individual interviews and the remaining five were interviews with more than one participant. Most interviews were conducted via Microsoft Teams (Microsoft Corp, Seattle 206 WA, USA) except for two interviews which were conducted in person, a group interview at the Department of Health in Pretoria, South Africa and one individual interview in London, UK. Regardless of the format of the interview, the responses from each participant were recorded on Slido (Cisco Systems., 2025) by a member of the research team. Slido is a website used to collect real time responses to closed and open text responses during live events, in our case an interview. For the group interview at the NDoH which had seven interviewees, responses were recorded in Slido by expertise area rather than individually (TB, EPI, and PHC). All interviewees were promised anonymity.

The interview guide was structured into five sections, and the questions asked to participants were dependant on their expertise and sector. Whilst most responses were qualitative in nature, a subset of questions generated quantitative data, as such the proportion of participants reporting a response to these questions has been reported accordingly. All participants, except those from regulatory or treasury/finance, were asked questions in section 1 on ‘Country Interest’ and section 2 on ‘Target Populations’ as a priority (supplementary file 1). Participants from the regulatory body were asked questions in section 3 on ‘Regulatory Considerations’ and those from finance/treasury were asked questions from section 6 on ‘Prioritisation of funding for TB vaccinations. Sections 4 and 5 were asked if time permitted but were less of a priority. For the section of the interview guide on target populations, participants were first asked to prioritise potential groups for vaccination, and follow-up questions were then limited to the participant’s top-priority group(s). Due to the flexible nature of the interviews, not all questions within each section were asked consistently across participant depending on the flow of the interview and time constraints, as such there is variation in the denominators of quantitative data reported in the results.

### Ethical considerations

All participants gave informed consent prior to the interviews. For participants who were interviewed over Microsoft Teams they were sent a participant information sheet and consent form over email in which they had to digitally sign their signature and send it back to us. For participants who were interviewed face-to-face they were provided with a paper copy of the participant information sheet and consent form which they signed and gave back to us. The study was approved by the London School of Hygiene & Tropical Medicine Ethics Committee (31511 /RR/37396) and the University of the Witwatersrand Johannesburg Human Research Ethics Committee (250102).

### Analysis

We conducted a deductive analysis, using predefined themes based on the interview guide (timelines for TB vaccine introduction, target populations, regulatory considerations, financing, and lessons from existing health programmes targeting adolescents and adults). Data collected via Slido during the interviews were exported into Microsoft Excel (Microsoft Corp, Seattle 206 WA, USA) for analysis. Data were formatted so each row of excel represented one participant’s data. We used a manual framework approach [18] for the analysis in which we generated codes from the themes in the interview guide, examples of derived themes were ‘efficacy’ and ‘price’. These codes were then grouped together into themes to form our analytical framework, in which a separate spreadsheet was created for each theme. Each row of data was then carefully read and indexed using the codes and relevant responses were sorted and entered line-by-line into the appropriate thematic spreadsheet to enable detailed review. Following this the data were then charted and an analytical memo was written for each of the themes to document the key thoughts, interpretations and reflections of the data, these were discussed amongst the team as a means of triangulation. Data were reported by expertise of participant rather than individually to ensure confidentiality and privacy of the participant.

Interview questions that were closed or clearly defined were analysed using descriptive quantitative methods and supplemented with qualitative comments where provided. Implementation strategies for the prioritised target populations were summarised in a table documenting the proposed approach for each target group, the expected coverage of a TB vaccination and timeline to achieve this.

## Results

A total of 26 interviewees participated in this study. Their expertise included TB policy and research, civil society/ TB advocacy, employment in the NDoH, regulatory oversight, National Treasury, and academia. An overview of interviewee expertise is summarized in supplementary Table 1.

### Section 1: Country interest

Stakeholders expressed widespread support for introducing a new TB vaccine in South Africa, driven largely by the country’s high burden of disease. However, they noted that several factors would shape the scale of adoption and roll-out pattern for new TB vaccines, as summarized in Figure 1.

**Figure 1:**
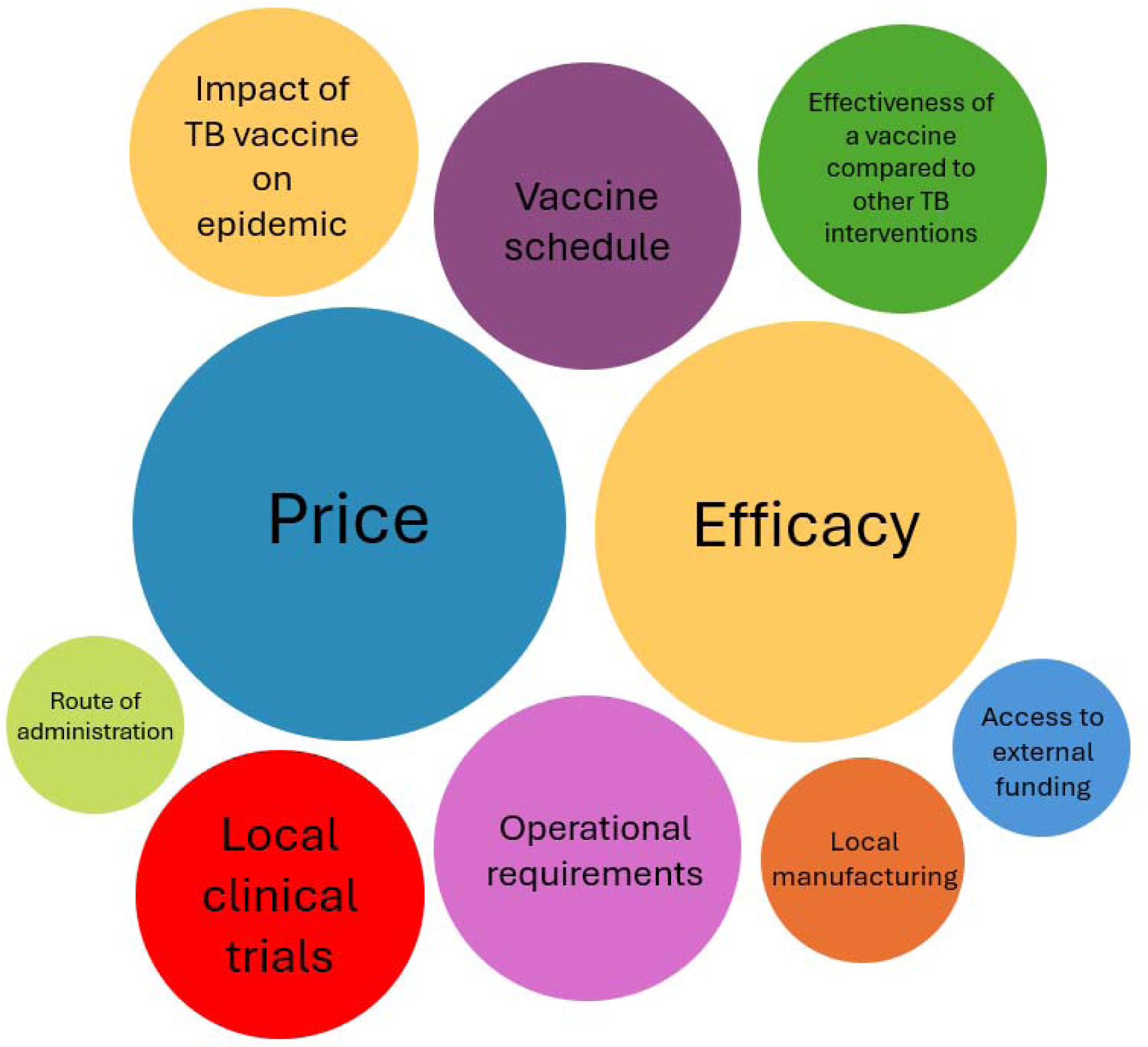
Factors influencing TB vaccine introduction.

Price was mentioned by the largest number of stakeholders (n=18/25) as being an important factor when considering the feasibility, scale, and speed of vaccine introduction. Stakeholders suggested a cheaper vaccine per course would be preferable as it would likely increase the availability of a vaccine, whereas a more expensive could delay introduction or restrict implementation to priority high-risk groups. The importance of modelling to understand the cost-effectiveness of TB vaccines compared to existing prevention measures such as TB preventive treatment was emphasized, with respondents noting that the National Treasury requires formal economic evaluations and investment cases that incorporate not only direct costs, but also indirect costs and savings from averted treatment courses, lost income, and reduced transmission. In addition to understanding whether a new TB vaccine was cost effective, the affordability of the vaccine within the current fiscal environment in South Africa was also seen as critical.

Vaccine efficacy was another key consideration for the roll-out of a vaccine and was mentioned by the majority of interviewees (n=17/25). WHO guidance suggests that an adult and adolescent TB vaccine should ideally demonstrate at least 50% efficacy [14], and interviewees expressed mixed views on whether implementation would proceed if efficacy fell below this threshold. Whilst some stakeholders suggested the vaccine would be introduced with lower than 50% efficacy, this was only said to be feasible if there was robust modelling and cost-effective analyses that demonstrated there was still a significant public health impact. Others suggested that introducing a vaccine with lower than 50% efficacy had the potential to undermine public trust and national policy credibility, which was especially important in the post-COVID-19 context. Participants reflected on the balance between individual-level efficacy and population-level impact, even with modest efficacy. If the vaccine could be shown by modelling to reduce TB incidence and mortality at scale, it would likely be acceptable to policymakers.

Stakeholders suggested interferon gamma release assay (IGRA) testing would not be feasible prior to vaccination in the general population and so demonstrating efficacy among IGRA negative populations was also a key consideration. However, given familiarity with the clinical trial design, stakeholders highlighted the possibility of off-label recommendation for TB vaccine use in IGRA negative populations and people living with HIV (PLHIV) in the absence of efficacy data for these groups in the initial introduction years. Similar to price, vaccine efficacy was suggested to heavily influence the implementation strategy, with lower efficacy resulting in a more targeted approach prioritizing high-risk groups rather than a broad population-based approach. Whilst vaccine efficacy was of great importance, decisions on the minimum efficacy of a vaccine were said to be heavily dependent on other factors such as cost-effectiveness, safety and feasibility of the delivery, and any decision that is made would need to be strongly informed by available evidence.

Participants were also asked about how important the vaccination schedule of a potential new TB vaccine would be; the majority (n=12/17) suggested it would be rolled out whether it was a one or two dose vaccine. However, participants strongly emphasized that a simple and effective dosing schedule is critical for vaccine uptake. Single-dose vaccines were consistently preferred by stakeholders due to ease of delivery, reduced logistical burden, and better adherence, particularly when reflecting on the poor uptake of second doses during COVID-19. Additionally, most of the participants stated they thought the vaccine would be introduced whether it was intramuscular or intradermal (n=12/16). However, discussions highlighted that participants expressed a preference for intramuscular administration over intradermal due to its ease, familiarity, and lower training burden. While intradermal was acknowledged as a viable option, particularly if efficacy was higher, it was considered more challenging due to concerns about side effects and the need for additional health worker training. Route of administration wasn’t seen as the most critical factor for TB vaccine introduction, but stakeholders noted intramuscular would be prioritized if both routes were similarly effective.

Participants also discussed how vaccination fits within the broader landscape of TB prevention strategies, particularly in relation to tuberculosis preventive therapy (TPT). TPT and vaccinations were consistently viewed as complementary strategies but with vaccines taking a priority (n=9/20). The balance between them was said to be dependent on efficacy, cost-effectiveness, and operational feasibility. Stakeholders generally preferred vaccines over TPT due to their perceived greater acceptability, simpler delivery, and potential for higher uptake, particularly if formulated as a single-dose vaccine. Many respondents noted that uptake of TPT remains low in South Africa due to challenges such as pill burden, multiple visits, and patient adherence, barriers that a vaccine may overcome.

South African Health Products Regulatory Authority (SAHPRA) requires local safety and efficacy data for TB vaccine registration, underscoring the importance of evidence tailored to the local population. South Africa frequently participates in, and often leads, global TB vaccine trials, ensuring that data reflect the local epidemiology. While local manufacturing of TB vaccines is viewed as preferable, it is not deemed essential for vaccine adoption. However, domestic manufacturing could influence procurement strategies, pricing, and long-term sustainability by providing greater supply certainty, enhancing political support, and strengthening local health system capacity. Drawing lessons from COVID-19, stakeholders recognized the value of building local manufacturing capabilities but acknowledged high initial costs and accepted that there may be a need to leverage existing partnerships with manufacturers internationally. Ultimately, it was thought, decisions around local manufacturing will depend on cost-effectiveness, quality assurance, and sustainability.

### Section 2: Target populations and introduction timelines

There was consensus that a broad country-wide population-based TB vaccination approach would be ideal given that TB occurs outside defined high-risk groups, but realistically a targeted strategy focusing on high-risk groups (HRGs) may be more cost-effective and feasible initially. Given the uneven geographic distribution of TB, with four provinces accounting for the majority of TB incidence (Guateng, Eastern Cape, Western Cape, and KwaZulu-Natal), a phased, subnational roll-out starting with high-burden areas and HRGs was suggested by multiple stakeholders, including NDoH. Whilst some stakeholders expressed that targeting high burden areas or high-risk populations e.g. PLHIV, could lead to stigmatisation, the NDoH suggested this would be acceptable provided there is evidence of an increased burden in these populations/places. Overall, while a nationwide rollout is the goal to ensure equity and address population mobility, due to resource limitations and logistical challenges, stakeholders suggested beginning with targeted key populations and expanding broadly as funding and vaccine availability improve.

Stakeholders suggested that a TB vaccine introduction in South Africa could be relatively quick, with most participants stating it could start within two years of the vaccine being licensed, (n=13/21) and all participants expecting it to be rolled out within a maximum of five years. South Africa’s participation in clinical trials, its history of early adoption of TB interventions, and ongoing preparatory work were cited as enabling factors for early adoption of new TB vaccines. However, the speed of rollout was said to depend on prior groundwork, tender processes, financing, and vaccine hesitancy among the target populations. While some were optimistic due to strong advocacy from civil society, clinicians, and researchers and the possibility of fast-tracking regulatory approval, others expressed caution, noting that delivery planning, and procurement and tender processes can take years unless exceptional measures are taken.

Stakeholders identified several key populations as priorities for a potential new TB vaccine rollout in South Africa. Their selection was shaped by considerations of disease burden, feasibility of vaccine delivery, and anticipated levels of acceptability. The most frequently mentioned groups were adolescents, PLHIV, adults, healthcare workers (HCWs) and TB household contacts. Additional populations that were mentioned were prisoners, miners, and people living with diabetes.

Adolescents were commonly cited as a primary target group due to their perceived high TB burden and transmission. Stakeholders highlighted the existing infrastructure for school-based vaccine delivery as an advantage, referencing the successful human papillomavirus (HPV) vaccination programme, which has achieved high coverage of over 70% [19]. Based on this, participants reported they expected a coverage for adolescents for TB vaccines to be between 50-90%, and that this coverage could be achieved within 5 years (Table 1). Adolescents in school were seen as both accessible and likely to accept the vaccine, though those out of school will be harder to reach.

**Table 1:** Suggested implementation strategies for the priority populations.

| Target population | Geographical scope | Delivery strategy | Coverage target | Years to achieve coverage | Feasibility |
| --- | --- | --- | --- | --- | --- |
| Adolescents (12–17-year-olds) | Nationwide most reported; phased subnational among high burden provinces roll out also suggested by several stakeholders | Campaign most reported; routine immunization also reported.<br>Most stakeholders suggested the two would run in parallel | Campaign<sup>1</sup> = 50-80%<br>Routine<sup>2</sup> = 60%-90% | Campaign and routine immunization majority of responses said between 1-5 years | Can reach population through schools, majority of adolescents are in school<br>HPV vaccination in schools has been successful |
| People living with HIV | Nationwide most commonly reported, regional also suggested | Both campaign and routine immunization equally reported | Campaign = 40-70%<br>Routine = 40-90% | Campaign = 1-5 years<br>Routine immunization = Most reported 3-5 years, one suggested 1-2 years | Easy to reach those who are diagnosed through HIV clinics for ARV collection & annual blood tests |
| Health care workers | Nationwide | Campaign most reported, routine immunization also reported. | Campaign = 80%<br>Routine = 50-80% | Campaign = 3-5 years<br>Routine immunization = 3-5 years | Easy to reach through workplace |
| Adults (18-64) | Nationwide most reported, regional also reported. | Campaign most reported; routine immunization also suggested<br>Most stakeholders suggested the two would run in parallel | Campaign = 50-60%<br>Routine 60-70% | Campaign = 1-10 years<br>Routine immunization = 1-10 years | A hard-to-reach population due to no existing infrastructure that vaccinates adults |
| Household contacts of TB | Nationwide | Campaign approach | 50% |  | Could be reached through contact tracing, although the logistics would be challenging |

<sup>1</sup>Campaign: vaccinate all participants within the group
<sup>2</sup>Routine immunization: vaccinate new entrants to the group

PLHIV were widely recognized as a high-priority group due to their increased risk of TB disease. Among those already diagnosed and engaged in care, vaccine delivery was considered feasible, with stakeholders suggesting integration into existing touch points with HIV patients such as annual viral load monitoring or antiretroviral therapy (ART) collection. Stakeholders suggested coverage among PLHIV would be between 40-90% with most suggesting it would take between 3-5 years to achieve this (Table 1). PLHIV were viewed as both accessible and likely to accept vaccination, particularly given their frequent contact with the health system. South Africa also has a high proportion of all PLHIV who are on ART at 77% which would minimise the concern around vaccinating those who are immunocompromised [20].

Although adults represent a population highly affected by TB, stakeholders described them as among the most difficult to reach. As such the anticipated vaccination coverage target among adults was suggested to be between 50-70% which would be achieved anywhere between 1-10 years (Table 1). Many healthy adults do not routinely engage with the health system and may demonstrate low health-seeking behaviour. Additionally, there is no existing adult vaccination infrastructure in South Africa, posing logistical challenges. Acceptability was perceived as variable and potentially low, particularly among healthy adults who do not see themselves as at risk.

Healthcare workers were described as a high-risk group due to occupational exposure, including among those not directly working in TB services. However, stakeholders noted a generally hesitant attitude toward vaccination within this group, which may present a barrier to uptake. Nevertheless, HCWs were also seen as influential actors in promoting vaccine confidence among the general population, making their inclusion strategically important for both reducing transmission within healthcare setting and promoting uptake within the broader community. Due to being a contained population, a nationwide roll out was suggested for this priority group with stakeholders suggesting a vaccination coverage target between 50-80% which could be achieved between 3-5 years (Table 1).

Some stakeholders mentioned TB household contacts as a potential target group, given their elevated risk of infection. While theoretically accessible through existing contact tracing programmes, this approach was viewed as operationally challenging and resource intensive. As such, a vaccination coverage target of 50% was suggested by participants (Table 1). Acceptability was expected to be higher within this group due to personal or familial experience with TB, but actual reach and feasibility were seen as limited.

### Section 3: Other health programs

Learning from previous vaccination and health programs targeting adolescents and adults, vaccine hesitancy was commonly referred to by stakeholders as a concern which necessitates strong communication and public education to build trust, particularly around the long-term benefits of TB vaccination. Interviewees mentioned that public messaging should emphasize the collective value of high vaccine uptake, including among individuals without direct experience of TB, to achieve population-level immunity and reduce overall transmission. Stakeholders also noted the lack of robust surveillance systems for vaccination documentation, highlighting the need for integrated, accessible systems with strengthened capacity at provincial and district levels. Effective rollout was said to require close collaboration with healthcare and community workers, using relatable storytelling, especially by TB survivors and targeted information campaigns through diverse media and civil society partners. Understanding community demand was also expected to be critical to guide successful implementation. An additional consideration raised by the NDoH was the limited number of nurses, of which are the only healthcare workers able to vaccinate.

### Section 4: Regulatory considerations

Decisions on off-label vaccine use are typically made by the NDoH, with the National Advisory Group on Immunization (NAGI) providing evidence-based recommendations. Off-label use may be approved in specific cases when safety concerns are minimal, as seen during the COVID-19 rollout. The Section 21 consent process allows clinicians to administer vaccines off-label with patient approval; however, it was suggested that this would not be ideal for large-scale campaigns. There was a consensus among stakeholders that IGRA screening would be not implemented as a pre-requisite for receiving the vaccine, provided that safety data for IGRA negative populations was available. Vaccination of PLHIV was considered safe for people whose disease was well controlled, indicated by a higher CD4 count, but could be a concern for those with a low CD4 count. Stakeholders also expressed a preference for multidose vials over single-dose vials and pre-filled syringes, citing lower cost, reduced storage volume, and more efficient use of cold chain storage as key advantages. Regarding timelines, participants from SAHPRA suggested new TB vaccines will likely be eligible for fast-track regulatory approval which could reduce review time to about 180 business days, compared to the standard 350 business days.

## Discussion

We conducted interviews with stakeholders in South Africa to understand their interest, priorities, and preferred rollout approach for a potential new TB vaccine for adolescents and adults. Building on previous literature, our findings reinforce the importance of early planning prior to the introduction of a new TB vaccine to ensure there are clear delivery strategies and an understanding of the logistical requirements for implementation [21, 22].

Overall, stakeholders from a broad range of expertise were supportive of a potential new TB vaccination due to the high burden of TB. However, they stated the implementation would be guided by several factors including efficacy, cost effectiveness, regulatory and logistical considerations. Price was mentioned as an important factor for vaccine introduction by the majority of stakeholders, who suggested a cheaper vaccine would be preferrable to increase availability and ensure a broad population-based implementation. Efficacy was noted as a core consideration, with a threshold below 50% seen by some stakeholders as a concern unless it was supported by robust modelling that demonstrated a significant public health impact and cost-effectiveness. Stakeholders mentioned a preference for a single dose vaccine that was administered intramuscularly, although these factors were not seen as essential prerequisites. There was also a preference for the vaccine to be a multi-dose vial as opposed to pre-filled syringes to reduce costs and storage requirements. A new TB vaccine is likely to qualify for fast-track regulatory approval process, which would take around 180 business days to be approved by SAPHRA. While local manufacturing was preferred as it was seen to enhance capacity and ensure supply, manufacturing in a country with which South Africa has an existing manufacturing partnership was also considered acceptable. Stakeholders indicated that the ideal implementation strategy was to introduce the vaccine to the general population, but if there were financial or logistical constraints, a more realistic approach would be to target key high-risk populations of which adolescents, PLHIV, HCWs were cited as the highest priority groups.

The suggested target populations were aligned with the local epidemiology and demographics of South Africa as well as considering the feasibility of vaccinating these target populations [5]. Adolescents are known to be a population that has a large proportion of incident TB cases and deaths and contribute to a considerable amount of TB transmission [5]. In addition, stakeholders see them as an accessible target population due to the ability to adapt the existing vaccination framework for HPV in schools [13, 23]. The prioritization of PLHIV is unsurprising given the high prevalence of HIV in South Africa, the fact they have one of the highest HIV and TB coinfection rates, and the poor TB outcomes among PLHIV [24]. HCWs were seen as a key population by stakeholders, not only due to their increased exposure to TB, but also given their influential role in patient education and changing attitudes and behaviors. In addition, HCWs were perceived as a group with higher vaccine hesitancy, a trend well-documented since COVID-19, where concerns about side effects, long-term health implications, and perceived lack of personal benefit contributed to declining uptake rates [25]. Stakeholders suggested that a combination of campaign and routine vaccination strategies would be necessary for a successful rollout, emphasizing that the availability of existing infrastructure, such as schools for adolescents and HIV clinics for PLHIV would be a critical determinant of feasibility [13].

In terms of the operational feasibility, stakeholders strongly favoured a single-dose schedule due to easier administration which would likely improve coverage and reduce burden on health systems. This was consistent with prior research which suggested a two-dose vaccination was seen by stakeholders as a potential barrier to uptake, most notably due to loss to follow up which has been seen previously with both COVID-19 and HPV [13, 17]. In addition to number of doses, depending on how rapidly vaccine-induced immunity declines, the frequency of doses would also likely influence the success of a multiple dose vaccine. Intramuscular administration was preferred over intradermal due to its simplicity, familiarity among HCWs, and lower training requirements, though intradermal delivery was seen as acceptable if efficacy benefits could justify its use. These considerations of schedule and route of administration will be important in the future, as leading candidates have conflicting characteristics. While M72, the most advanced TB vaccine currently in Phase III trials, is administered intramuscularly, its two-dose regimen may pose logistical challenges. In contrast, MTBVAC, which is in Phase IIb trials, offers the advantage of a single-dose schedule but requires intradermal administration, which may complicate delivery due to training and operational demands [10].

The potential for local manufacturing was also discussed as a strategic opportunity. Although not essential, domestic production was seen as a long-term enabler of supply security, affordability, and political support. Lessons from COVID-19 highlighted the value of building local capacity, though stakeholders acknowledged the high upfront costs and emphasized that sustainability, quality assurance, and cost-effectiveness would ultimately determine feasibility of local vaccine production.

Regarding implementation feasibility, stakeholders noted the limited number of nurses available to administer vaccinations which was presented as a key barrier for nationwide implementation. This has also been highlighted in a previous research conducted in South Africa but also more broadly in low- and middle-income countries [13, 17].

Our study highlighted that local clinical trials were important to stakeholders as a means to provide evidence specific to the local population and promote confidence among policy makers and the public [26]. Following licensure, local manufacturing was seen as beneficial to ensure adequate supply and to reduce the costs in the long term, however stakeholders were aware that that new vaccine technologies usually have a higher initial market price which can make investment cases harder to justify [21].

Routine IGRA testing prior to vaccination was considered neither feasible nor necessary, provided sufficient safety data in IGRA-negative populations was available, this was due to the costs and lack of feasibility, which is particularly important given the current uncertainty around vaccine efficacy in IGRA-negative individuals [12, 13].

Stakeholders expected that the SAHPRA would likely support accelerated review pathways, particularly given the country’s active participation in TB vaccine trials and the public health urgency. Decisions around off-label use were expected to follow established pathways for approval. Overall, the regulatory environment was viewed as encouraging to rapid implementation, contingent on strong evidence and coordinated engagement among government, researchers, and manufacturers.

A barrier raised frequently by stakeholders was the concern around vaccine hesitancy, which, whilst already well documented has risen since COVID-19 and poses a threat to vaccine uptake for a new TB vaccine, particularly if the public does not perceive themselves to be at risk [17, 27–29]. Additionally, stakeholders also raised concerns around increased vaccine hesitancy if the efficacy of a new TB vaccine were low. This concern is supported by COVID-19 research, which identified three main drivers of vaccine hesitancy: perceived disease risk, vaccine safety, and vaccine efficacy [30]. Research conducted in Mozambique highlighted that hesitancy of a TB vaccine was higher for new vaccine than for a booster of an existing one e.eg BCG, suggesting that educating the public on a new vaccine will be essential [31]. This is in line with both previous research on TB vaccines among stakeholders in South Africa and our findings which emphasized the need for increased civil society engagement to increase education, promote vaccine uptake and build trust among the public ahead of licensure [4].

This study addresses a key evidence gap in the implementation of a new TB vaccine in South Africa which is important and timely given the ongoing clinical trials to develop a safe and efficacious TB vaccine. The use of semi-structured interviews provided flexibility, allowing discussions to be tailored to individual participants’ expertise and experiences. However, it had several limitations. First, it did not include interviews with the general population or those who are likely to, which limits insight into community-level perspectives and vaccine acceptability. Additionally, some data were collected via group discussions and recorded on Slido on behalf of multiple interviewees, which may affect the richness of individual responses. Certain stakeholder groups may have been underrepresented, and findings are not generalizable beyond the South African context due to demographic and epidemiological differences. The use of Microsoft Teams for interviews, while necessary, may have impacted participant engagement and data quality [32]. Finally, as the vaccine is not yet available, participants responded to hypothetical scenarios, leaving many uncertainties regarding actual vaccine price, population willingness to vaccinate, and real-world vaccine efficacy. These factors highlight the need for ongoing research as more empirical data become available.

Moving forward, it is essential to understand the acceptability of receiving a new TB vaccine in the priority populations such as adolescents, PLHIV and HCWs. While vaccinating these groups has potential for a large public health impact, low uptake of the vaccine due to hesitancy and poor acceptability of the vaccine, may limit the impact. Similarly, while our findings highlight that pre-vaccination infection status testing is unlikely to be conducted, some vaccines may only be effective in those individuals who are already infected with *Mtb*. Therefore, it is important to understand the TB infection prevalence in these populations to allow for better modelled estimates of the potential impact of introducing a new TB vaccine. Further research is needed to collect empirical data on vaccine acceptability and TB infection prevalence to model the estimated cost effectiveness and budget impact.

## Conclusion

In South Africa stakeholders were very supportive of a new TB vaccine for adolescents and adults due to the high burden of TB but highlighted several factors that would influence the implementation. A targeted approach focusing on high-risk groups such as adolescents, PLHIV and HCWs was seen to be most likely in anticipation of limited resources and funding. We hope this work will inform the National Advisory Group on Immunisation (NAGI) and support its submission of a vaccine recommendation to the NDoH in South Africa. It will also support further preparatory activities being conducted in South Africa such as the WHO and NDoH stakeholder engagement workshop in July 2025.

Further research should focus on collecting empirical data on vaccine acceptability and TB infection prevalence, alongside modelling to estimate cost-effectiveness and budget impact, to support informed decision-making and maximise public health benefits.

## Data Availability

We cannot share the dataset as we ensured confidentiality to all of our interviewees, and the data could be linked to them. As such, we are unable to provide the anonymised dataset. However, we are happy to share the data upon request to interested, qualified researchers. Please contact Katherine Thomas or Richard White (PI and co-last author). Data will be stored on a secure server for 2 years, and researcher notes will be stored in a locked cabinet. Data will be managed by the project coordinator and will only be accessible by the study team, under the guidance of the PI. Interested researchers will be asked to complete a data sharing agreement on request for access to the anonymised data. Although the authors cannot make their study’s data publicly available at the time of publication, all authors commit to make the data underlying the findings described in this study fully available without restriction to those who request the data.

## Acknowledgements

We thank all the anonymous interviewees.

We also thank Julio Croda, Marcio Natividade, Layana Alves, Prof Sri, Nina Dwi Putri, Puck Pelzer, Heidi Larson, Rupali Limaye, Joeri Buis, Andrew Kerkhoff, Kristin Nelson, Jody Boffa, Zsofia Hesketh, Crhistinne Gonçalves, Ezio Tavora Dos Santos Filho, Lisa Cranmer, Rudzani Muloiwa, Ben Kagina, Frauke Uekermann, Tara Lavanya Prasad, Tiziana Scarna, Adam Soble, Birgitte Giersing, Forum Mistry and Sarah Ewart who helped design the questionnaire.

## Notes

### Competing Interest Statement

KAT, CO, TN MK,VD, DG, OR, RAC, WJ, FA, SC, JS and RGW have no competing interests. GJC is funded by the Gates Foundation through a grant to the Aurum Institute to implement a first-in-human TB vaccine trial. GJC is the Chair of the WHO Technical Advisory Group on Evidence for Clinical and Policy Considerations for New Tuberculosis (TB) Vaccines.

### Funding Statement

This work was funded by Wellcome Trust (310728/Z/24/Z, 218261/Z/19/Z). KAT is funded by Wellcome Trust (310728/Z/24/Z). RAC was funded by Open Philanthropy/Good Ventures (GV673606227), Wellcome Trust (310728/Z/24/Z), BMGF (INV-001754) and NIH (G-202303-69963, R-202309-71190) to LSHTM. RGW is also funded by the Wellcome Trust (310728/Z/24/Z, 218261/Z/19/Z), NIH (1R01AI147321-01, G-202303-69963, R-202309-71190), EDTCP (RIA208D-2505B), UK MRC (CCF17-7779 via SET Bloomsbury), ESRC (ES/P008011/1), BMGF (INV-004737, INV-035506), Open Philanthropy/Good Ventures (GV673606227), and the WHO (2020/985800-0). Co-authors at CHAI (CO, VD, DG & OR) received funding from the Gates Foundation (INV-059503).

### Author Declarations

This study was approved by the London School of Hygiene & Tropical Medicine Ethics Committee and the University of the Witwatersrand Johannesburg Human Research Ethics Committee

### Summary of Updates

This version of the manuscript has been revised to include the correct dates of which participants were contacted to be included in the study

