## Supplemental File 1 for "Insights on the Implementation Strategies for a Potential New Tuberculosis Vaccine in South Africa"

**Stakeholder Consultations for TB Vaccine Demand and Impact Modelling**

**Interview Guide**

**Legend:** Priority sections, questions, interviewer instructions, response

### **Instructions for the interviewer**

- Introduce yourself and the purpose of the interview (use ‘Introduction’ section below)
- Ensure the respondent understands that their individual responses will be kept confidential
- Read each question clearly and allow sufficient time for the respondent to answer
- Record the respondent's answers in detail
- Use the probes provided to elicit more detailed responses where necessary
- Specific instructions for the interviewer are in red texts
- Use the table below to tailor the questionnaire to each stakeholder prior to the interviews
- Prioritize sections 1, 2 & 3 during the interview and go through sections 4, 5 & 6 if time permits

| **Section** | **Relevant Stakeholders** | | | | |
| --- | --- | --- | --- | --- | --- |
|  | **MOH** | **National TB Program** | **EPI** | **Regulators/ NITAG** | **MOF** |
| 1. Country interest in TB vaccine introduction | X | X | X | X |  |
| 2. Indicative timelines and target population for TB vaccination | X | X | X |  |  |
| 3. Regulatory considerations |  |  |  | X |  |
| 4. Experience from other vaccines or health programs targeting adolescents and adults | X | X | X | X | X |
| 5. Tuberculosis/HIV control programs | X | X |  |  |  |
| 6. Prioritization of funding for TB vaccination (not required for initial round of consultations in some countries) | X | X | X |  | X |

### **General Information**

Before commencing each interview, complete the table below

| Country |  |
| --- | --- |
| Interview serial number (format 01, 05, 10, etc.) |  |
| Organization |  |
| Name of Interviewee(s) |  |
| Position(s) |  |
| Date of interview |  |
| Interviewer |  |
| Which of the following are your areas of experience?  Tick all relevant areas | ☐ Tuberculosis prevention, diagnosis & treatment  ☐ Vaccine development  ☐ Vaccine procurement/supply  ☐ Vaccine delivery  ☐ Health/programme financing  ☐ Regional/local level  ☐ National level  ☐ Regulatory bodies  ☐ Bodies for the incorporation of technologies in the public health system  ☐ NGO of affected population  ☐ Policy maker e.g., NITAG  ☐ Other (please specify): __________________ |

### **Introduction**

Several TB vaccine candidates in late-stage clinical development could have a transformative impact on adult and adolescent populations when they come to market over the next decade. Early market-shaping efforts are already underway to accelerate access and avoid delays like those experienced with some previous vaccine roll-outs, e.g., malaria.

As part of these efforts, CHAI and LSHTM are conducting consultations with high-level country stakeholders to improving demand visibility for TB vaccines. Stakeholders being consulted include those responsible for strategic decision making on policy, regulatory aspects, financing, procurement and delivery in relation to future TB vaccines. We will use the insights generated from the consultations to develop realistic demand scenarios for future TB vaccines. The demand scenarios will be shared with stakeholders in the TB vaccine ecosystem to inform decisions on further R&D, supply planning, global policy and financing.

This interview will last for about one hour and has X sections

Section 1: Country interest in TB vaccine introduction

Section 2: Indicative timelines and target population for TB vaccination

Section 3: Regulatory considerations

Section 4: Experience from other vaccines or health programs targeting adolescents and adults

Section 5: Tuberculosis/ HIV control programs

Section 6: Prioritization of funding for TB vaccination

In this interview, we would like to gain a detailed understanding of your country’s potential approach to TB vaccination – assuming unlimited funding and supply – based on public health needs. In later discussions, we can explore the potential implications of budgetary constraints on implementing a TB vaccine program in your country.

All insights generated from the consultations will be aggregated and anonymized. Individual insights will remain confidential to our project team.

Pause for any questions from the respondent and confirm if the interview can be recorded to ensure that insights are accurately captured.

Can we contact you later if we run out of time/have follow-up questions?

☐ Yes

☐ No

Also confirm how best to do the follow-ups: xxx

### **Section 1: Country Interest in TB Vaccine Introduction**

1) If an efficacious and safe TB vaccine were available, do you think there would be interest in introducing it in your country?

Select one response and record the rationale

☐ Yes

☐ No

☐ Unsure

Rationale: xxx

*If ‘No’, record the rationale and end the interview*

2) Decision making on country willingness and timelines for TB vaccine introduction
   a) What are the main data points and factors that would influence your country’s decision and timeline to introduce TB vaccines?

Select all that apply, and record the rationale

☐ Vaccination schedule, i.e., one or two doses per fully immunized person

☐ Route of action of TB vaccines, i.e., intramuscular or intradermal

☐ Efficacy

☐ Effectiveness of TB vaccines compared to other TB interventions, e.g., TB preventive treatment

☐ Local manufacturing

☒ Local clinical trials

☐ Price

☐ Access to external funding

☐ Projected impact of TB vaccination on the TB epidemic

☐ Operational requirements e.g., storage temperature

☐ Others (specify below)

Rationale for each option selected: xxx

Probe with the following questions, if not already mentioned by the interviewee

i) How would the vaccination schedule impact your country’s decision on TB vaccine introduction?

Select one option and record the rationale

☐ Only introduce vaccine with one dose per fully immunized person

☐ Only introduce vaccine with two doses per fully immunized person

☐ Introduce whether one or two doses per fully immunized person

☐ Unsure

Rationale: xxx

ii) Would the route of action of the vaccines impact your country’s decision on TB vaccine introduction?

Select one option and record the rationale

☐ Only introduce intramuscular TB vaccines

☐ Only introduce Intradermal TB vaccines

☐ Introduce whether intramuscular or intradermal

☐ Unsure

Rationale: xxx

iii) Based on WHO’s guidance, efficacy of 50% or greater is preferred for TB vaccines. If efficacy was lower than 50%, would your country introduce the TB vaccine?

Select one response and record the rationale

☐ Yes

☐ No [If 2aiii = ‘No’, skip to 2av]

☐ Unsure

Rationale: xxx

[If 2aiii = ‘No’, skip]

iv) What is the minimum efficacy required for TB vaccines to be considered for introduction in your country?

Select one option and record the rationale

☐ 40%

☐ 30%

☐ 20%

☐ Less than 20%

☐ Unsure

Rationale: xxx

v) What other criteria (apart from efficacy) will your country use to compare the effectiveness of TB vaccines to other TB interventions, e.g., TB preventive treatment, to decide on whether to adopt the TB vaccines?

Record all answers from the interviewee

Response: xxx

vi) Would your country consider prioritizing TB vaccines over TB preventive therapy (TPT)/ other preventative measures?

Select one response

☐ Yes, TB vaccines would be prioritized over TPT

☐ Yes, TB vaccines would be prioritised over TPT, but only if the vaccine had certain characteristics (specify which characteristics in the rationale)

☐ No, TPT would be prioritised over TB vaccines

☐ No, TPT and vaccines would both be offered (specify how in the rationale)

☐ Unsure

Rationale: xxx

vii) What would be the main drivers of preferences between TPT and TB vaccines?

Select all that apply

☒ Price

☐ TB vaccine cost effectiveness compared to TPT

☐ Vaccine efficacy compared to TPT effectiveness

☐ Vaccine efficacy/safety in target populations compared to TPT

☐ Anticipated population acceptability (the population may prefer TB vaccines over TPT or vice versa)

☐ Unsure

More details/other drivers: xxx

[Skip if local manufacturing is not prerequisite for TB vaccine introduction based on question 2a]

viii) Does your country’s local manufacturing requirement imply that all components of the vaccines (i.e. the antigen and adjuvant) must be locally manufactured?

Select one response

☐ Yes, all components must be manufactured locally

☐ No, only some of the vaccine components must be manufactured locally

☐ Unsure

Rationale: xxx

[Skip if local clinical trial is not prerequisite for TB vaccine introduction based on question 2a]

ix) Will the country delay TB vaccine introduction until there are local clinical trials?

Select one response

☐ Yes, wait until there are local clinical trial results before introducing TB vaccines

☐ No, introduce TB vaccines, but wait with widespread scale-up until data from local trials is available

☐ No, introduce and scale up TB vaccines without local clinical trials

☐ Unsure

Rationale: xxx

[Skip if in question 2a ‘price’ is not selected]

For calibration, remind the interviewee of some of the cost per vaccinated individual for some of the vaccines used in your country.

| Vaccine | Vaccine cost per vaccinated individual |
| --- | --- |
| PCV | *[price in local currency]* |
| Rota | *[price in local currency]* |
| HPV | *[price in local currency]* |
| COVID-19 | *[price in local currency]* |
| … | *…* |

x) If the efficacy of a TB vaccine is 50% and this persists for 10 years, would the vaccines costs per vaccinated individual below, delay TB vaccine introduction? If so, how?

Select one response for each price

R$118/person	☐ Yes, would delay	☐ No, would not delay	☐ Unsure

R$59/person	☐ Yes, would delay	☐ No, would not delay	☐ Unsure

R$29/person	☐ Yes, would delay	☐ No, would not delay	☐ Unsure

R$15/person	☐ Yes, would delay	☐ No, would not delay	☐ Unsure

Rationale: xxx

b) What other barriers not already mentioned could hinder/ delay your country’s introduction of an adolescent/adult TB vaccine or limit its scale up, and recommended solutions?

Record responses using the table below, go through each of the barrier types to check for answers

| **Type of barrier** | **Description of the barrier** | **Potential implications**  On introduction timelines, phasing, scale of introduction, target population etc. | **Recommended solutions** |
| --- | --- | --- | --- |
| Political | Xxxxx | Xxxx | xxxx |
| Financing related | Xxxxx | Xxxx | xxxx |
| Regulatory | Xxxxx | Xxxxx | xxxx |
| Demand | Xxxxx | Xxxxx | xxxx |
| Others (specify) | Xxxxx | Xxxxx | xxxx |

### **Section 2: Indicative Timelines and Target Population for TB Vaccination**

3) Assuming TB vaccines are licensed, prequalified by WHO and available for purchase by 2030, when do you expect to introduce the vaccines?

For calibration, remind the interviewee of the introduction timelines for other vaccines. See some examples in the table below.

| Vaccine | Initially licensed in… | Recommended for worldwide adoption by WHO SAGE in… | Received WHO prequalification in… | Gavi launched in… | Brazil introduced in… |
| --- | --- | --- | --- | --- | --- |
| PCV | 2000 | 2006 | 2008 | 2009 | 2010 |
| Rota | 2006 | 2009 | 2008/2009 | 2011 | 2006 |
| HPV | 2006 | 2008 | 2009 | 2012 | 2014 |

Select one response and record the rationale

☐ Within 0–2 years

☐ Within 3–5 years

☐ Within 6–10 years

☐ Within 11–20 years

☐ After more than 20 years

☐ Unsure

Rationale: xxx

4) What would be your country’s preferred approach to determine the target population for TB vaccination?

Select all that apply

☐ Broad population-based approach e.g., adolescents, adults

☐ Risk group-based approach i.e., target specific high-risk groups e.g., PLHIV etc.

Rationale: xxx

5) Target population

Capture the interviewee’s responses using the tables below.

For the relevant table(s) under this question, first go through questions ‘a’ to ‘c’ to identify all the populations the country is interested in targeting for TB vaccination and their rationale. Then, ask question ‘d’ on ranking in order of importance for introduction.

i) Table 1: Fill this table if the respondent selected ‘broad population-based approach’ in question 4

| **Target population** | b) **Would your country want to introduce TB vaccines to this population?**   Select response | c) **Where in your country would you roll-out the TB vaccines to these population groups?** | **Rationale/ More details** | d) **If** **5a=‘yes’, please rank in order of importance for introduction e.g., 1 – most important, 2 - less important etc.** Record number |
| --- | --- | --- | --- | --- |
| Adolescents  (12–17 years) | ☐ Yes  ☐ No  ☐ Unsure | ☐ Nationwide  ☐ Specific subnational areas/ districts/ settlements (in which locations?) | XXX |  |
| Adults  (18–64 years) | ☐ Yes  ☐ No  ☐ Unsure | ☐ Nationwide  ☐ Specific subnational areas/ districts/ settlements (in which locations?) | XXX |  |

ii) Table 2: Fill this table if the respondent selected ‘risk group-based approach’ in question 4

For the rows labelled ‘Other’, read out these examples to the interviewee to confirm which additional populations the country might be interested in targeting: malnourished, people who have previously had TB disease, migrants, refugees, homeless people, people living in high-risk congregate settings, people with unhealthy alcohol use or substance misuse, people living in high-density areas, immunocompromised & immunosuppressed persons, pregnant women, lactating women, persons older than 65 years.

| **Target population** | a) **Would your country want to introduce TB vaccines to this population?**   Select response | b) **Where in your country would you roll-out the TB vaccines to these population groups?** | **Rationale/ More details** | c) **If** **5a=’yes’, please rank in order of importance for introduction e.g., 1 – most important, 2 - less important etc.** Record number |
| --- | --- | --- | --- | --- |
| People living with HIV | ☐ Yes  ☐ No  ☐ Unsure | ☐ Nationwide  ☐ Specific subnational areas/ districts/ settlements (in which locations?) | XXX |  |
| Household contacts of TB patients | ☐ Yes  ☐ No  ☐ Unsure | ☐ Nationwide  ☐ Specific subnational areas/ districts/ settlements (in which locations?) | XXX |  |
| Healthcare workers | ☐ Yes  ☐ No  ☐ Unsure | ☐ Nationwide  ☐ Specific subnational areas/ districts/ settlements (in which locations?) | XXX |  |
| Other *(Specify here. See examples in the text before this table)* | ☐ Yes  ☐ No | ☐ Nationwide  ☐ Specific subnational areas/ districts/ settlements (in which locations?) | XXX |  |
| Others *(Specify here. See examples in the text before this table)* | ☐ Yes  ☐ No | ☐ Nationwide  ☐ Specific subnational areas/ districts/ settlements (in which locations?) | XXX |  |
| Others *(Specify here. See examples in the text before this table)* | ☐ Yes  ☐ No | ☐ Nationwide  ☐ Specific subnational areas/ districts/ settlements (in which locations?) | XXX |  |

6) Delivery strategies and target coverage rates

Capture the interviewee’s responses using the table in the subsequent pages.

First review the target populations listed in the table and mark those to which the country plans to introduce TB vaccines, based on the response to question 5. Then, go through questions ‘a-h’ for the first target population of interest before moving to the next one.

For ‘c-f’ (expected coverage rates and timelines for achieving those), remind the interviewee about the current coverage rates for other vaccines in their country

| Vaccine | Coverage rate in Brazil (2023) | Years since introduction of the vaccine in Brazil |
| --- | --- | --- |
| PCV | 74% | 13 years |
| Rota | 87% | 17 years |
| HPV | XX% | 9 years |
| COVID-19 | 87% | 13 years |

| **Target population**  Tick and cover these detailed questions only if the interviewee selected ‘yes’ in question 5a | a) **What would be your country’s strategy for delivery of TB vaccines to this population?**   Select all that apply and record the applicable age ranges  Remind the respondent that the main age groups for TB vaccines will be adolescents & adults | b) **What is the reason for targeting this population with this delivery strategy?**   Record rationale | c) **For RI, what coverage rate would your country aim to achieve?**   Record expected coverage rates for RI where applicable  Skip if ‘RI’ was not selected in 6a | d) **How long after starting RI would you expect to reach this target RI coverage rate?**   Select one option  Skip if ‘RI’ was not selected in 6a | e) **For campaigns / catch-up, what coverage rate would your country aim to achieve?**   Record expected coverage rates for campaigns/ catch-up where applicable  Skip if ‘campaign/ catch-up’ was not selected in 6a | f) **How long after starting the campaigns / catch up would you expect to reach this target coverage?**   Skip if the respondent did not select ‘campaign/ catch-up’ in question ‘6a’ | g) **In what order will your country implement RI and campaigns/ catch-up?**   For each row, skip if the respondent did not select both ‘Routine immunization’ & ‘Campaigns’ in Question 6b | h) **Feasibility: Eg has the government been able to reach this population with other programs in the past?** |
| --- | --- | --- | --- | --- | --- | --- | --- | --- |
| Adolescents  (12–17 years)  ☐ | ☐ Routine immunization (RI) for age group: x-x years (specify)  ☐ Catch up campaigns/ immunization for age group: x-x years (specify)  ☐ Other (explain in the rationale) | Xxx | XX% | ☐ 1–2 years  ☐ 3–5 years  ☐ 6–10 years  ☐ >10 years  ☐ Other (specify): xxx | XX% | ☐ 1–2 years  ☐ 3–5 years  ☐ 6–10 years  ☐ >10 years  ☐ Other (specify): xxx | ☐ Implement RI first  ☐ Implement catch up campaigns first  ☐ Implement RI and catch up campaigns in parallel  More details: xxx | ☐ Yes  ☐ No  More details: xxx |

7) Pattern/ scale of introduction of TB vaccines
   a) Do you foresee a roll-out to the groups above, differentiated by geographies?

Select one response and record the rationale

☐ Roll out only in specific subnational areas/no nationwide roll-out

☐ Subnational roll-out first and nationwide scale-up later (in the rationale, record in which areas, and in what order of areas)

☐ Nationwide roll-out from the outset

☐ Unsure

Rationale: xxx

b) In the relevant geographies, would your country introduce to the target population(s) all at once?

Select one response and record the rationale, especially on the phasing and timing if they intend to phase the introductions

☐ Yes, introduce to the target population(s) all at once

☐ No, phase introductions to the target population(s)

☐ Unsure

Rationale:

c) Will supply or funding constraints change any aspect of your intended delivery strategies for the target populations/geographies?

☐ Yes

☐ No

If yes, how: xxx

### **Section 3: Regulatory Considerations**

8) Under what conditions does your country consider approvals for the off-label use of vaccines? I.e. administering vaccines for an indication, dosage, schedule, or population not specified in its approved labeling

Record all answers from the interviewee

Response: xxx

a) When the first adolescent and adult TB vaccines come to market (i.e., around 2030), safety data for people living with HIV and IGRA-negative individuals would be available, but likely no efficacy data for these groups yet. (IGRA is a test for TB infection)
   i) In the absence of efficacy data for IGRA negative people by 2030, do you anticipate the TB vaccines will be approved for use in IGRA negative populations in your country? Remind the interviewee that efficacy data for IGRA positive people should be available.

☐ Yes

☐ No

☐ Unsure

Rationale: xxx

[Skip, if 8ai = ‘yes’]

ii) Will IGRA pre-screening be a prerequisite for TB vaccination or would you skip IGRA screening?

Record in detail the answer from the interviewee.

☐ IGRA pre-screening will be a pre-requisite for TB vaccination

☐ IGRA pre-screening will not be a pre-requisite for TB vaccination

☐ Unsure

Rationale: xxx

[Skip, if 8aii = ‘yes’]

iii) What additional data will be required for a positive regulatory decision, i.e. approval for use in the IGRA-negative population in your country?

Record in detail the answer from the interviewee.

Response: xxx

[Skip if country will not prioritize PLHIV, based on question 5a]

iv) In the absence of efficacy data for PLHIV by 2030, do you anticipate the TB vaccines will be approved for use among people living with HIV in your country? Remind the respondent that efficacy data for HIV negative people should be available

☐ Yes

☐ No

☐ Unsure

Rationale: xxx

[Skip, if 8aiv = ‘yes’]

v) What additional data will be required for a positive regulatory decision, i.e. approval for use among people living with HIV in your country?

Record in detail the answer from the interviewee

Response: xxx

b) Would concerns of vaccinating individuals with compromised immunity with a live attenuated vaccine prevent your country from vaccinating these populations (e.g., PLHIV, people living with diabetes, the elderly or other individuals with comorbidities) with a live-attenuated TB vaccine? Remind the respondent that safety data for PLHIV will be available?

☐ Yes

☐ No

☐ Unsure

Rationale: xxx

9) We understand that in your country, the registration process for vaccines is highly variable and could take a few months to a 3 years. Do you expect the same timeline for registration of TB vaccines, or do you see the option to fast track?

☐ Regular registration process/timelines

☐ Fast-tracked registration process

☐ Unsure

Rationale: xxx

Probe using the questions below if not already mentioned

a) Are interventions for TB control classified as public health priorities that qualify for fast-tracked regulatory approvals like COVID-19 vaccines?

☐ Yes

☐ No

☐ Unsure

More details: xxx

b) What is the timeline for fast-tracked approvals compared to the normal process for regulatory approval?

Timeline for fast tracked approval: X business days

More details: xxx

c) For accelerated access to TB vaccines, what other requirements not already discussed are critical from the regulatory perspective and what information will be required for evaluation of these requirements in your country?

Record all answers from the interviewee

More details: xxx

### **4:** **Experience from other Vaccines or Health Programs**

Recall that the main age group for TB vaccines will be adolescents and adults. In this section, we will explore lessons learned from your experience implementing other vaccination/ health programs among these age groups for example COVID-19, HPV & Tetanus Toxoid vaccination, sexual & reproductive health programs, TB prevention and treatment programs etc.

10) Could you share key lessons from other vaccination or health programs that have targeted similar age groups?

a) What went well?

Record all answers from the interviewee

Response: xxx

b) What were some of the challenges?

Record all answers from the interviewee

Response: xxx

c) How can these lessons be applied to the TB vaccination program?

Record all answers from the interviewee

Response: xxx

d) What was done to build demand for these vaccines/health programs (e.g., advertisements, campaigning etc.)?

Record all answers from the interviewee

Response: xxx

e) What coverage was achieved? How long did it take to achieve this coverage?

Record all answers from the interviewee

Response: xxx

f) Can you describe some of the logistical details of how the vaccine was introduced and delivered?

Record all answers from the interviewee

Response: xxx

g) Which budgets did these come from (i.e., who was the payer for the different items)?

Record all answers from the interviewee

Response: xxx

11) What infrastructure, systems, processes, resources etc. from these programs could be leveraged for the TB vaccination program?

Record all answers from the interviewee; probe on existing technical advisory/ decision making/ coordination groups, data review/management platforms, fast-track regulatory processes, cold chain equipment, communication systems etc.

Response: xxx

**Section 5: Tuberculosis/HIV Control Programs**

Only for MOH/ participants with TB experience

12) Do you think there will be changes in the TB control programme in the future (e.g. changes to treatment regimens or other control measures)?

☐ Yes

☐ No

☐ Unsure

If yes, what do you think will change, how do you think it will change and when?: xxx

13) Do you think there will be changes in the HIV control programme in the future (e.g. changes to treatment regimens or other control measures)?

☐ Yes

☐ No

☐ Unsure

If yes, what do you think will change, how do you think it will change and when?: xxx

14) How do you think coordination between TB and vaccine programmes will work on policy, funding and execution?

Record all answers from the interviewee

Response: xxx

15) Tuberculosis preventive therapy (TPT) experience. Skip if no TPT experience in this country
    a) What is the experience in your country with other TB prevention measures, particularly tuberculosis preventive therapy (TPT), and can you describe some of the logistical details of how these were introduced and delivered?

Record all answers from the interviewee

Response: xxx

b) Can you describe the costs of the resources and services you just described? If you do not know the cost, do you know where this data could be found?

Record all answers from the interviewee

Response: xxx

16) How should TB vaccination be integrated with other preventative measures such as tuberculosis preventive therapy (TPT)?

Record all answers from the interviewee

Response: xxx

**Section 6: Prioritisation of Financing for TB Vaccination**

Only for participants with health/programme financing experience

17) Is there, or will there be, an anticipated financing mechanism available to support the introduction of a new TB vaccine in your country?

☐ Yes

☐ No

☐ Unsure

More details: xxx

18) Are you able to share the approximate governmental budget for the TB programme, the immunisation programme, and/or the HIV programme? If you do not know the budget, do you know where this data could be found?

Record all answers from the interviewee

Response:

TB program: xxx

Immunisation program: xxx

HIV program: xxx

19) Who would need to give approval for reallocation of existing funding or allocation of new funding to cover the resource requirements of this program? What do you think they would consider in making this decision?

Record all answers from the interviewee

Response: xxx

a) What are some of the ways to expand budget allocation to TB & Immunization programs to scale up the vaccine to cover all eligible population? (like Health & Education Cess levied by the Indian government)

Record all answers from the interviewee

Response: xxx

b) Who are the key stakeholders responsible for such a decision?

Record all answers from the interviewee

Response: xxx

c) What is the evidence required to make such a case?

Record all answers from the interviewee

Response: xxx

20) When thinking about a potential budget for a TB vaccine, do you consider impacts on other areas beyond the direct costs and benefits of this program when thinking about how much to spend?

*For example:*

- *Reduced treatment costs of recipients in the future*
- *Impacts on other disease areas e.g. if someone living with HIV is prevented from dying from TB, then they will be treated for HIV for longer*
- *Impacts on other members of the family of the treated patient e.g. financial impoverishment, caring time and effort etc.*
- *Impacts on the wider economy in terms of the economic contribution of a healthy population*
- *Any other impacts?)*

Record all answers from the interviewee

Response: xxx

21) If estimates of the economic impact of introducing new TB vaccines are available, who would we best share those with?

Record all answers from the interviewee

Response: xxx

### **Wrap up/ closing**

Is there anything else you would like to add?

Thank you for your time.
