## Supplemental Table 1 for "Insights on the Implementation Strategies for a Potential New Tuberculosis Vaccine in South Africa"

| **S1 Table: Overview of expertise of interviewees** | |
| --- | --- |
| **Area of expertise** | **Number of participants** |
| Researcher | 4 |
| NGO/ Researcher | 4 |
| Global organization/ funder | 2 |
| Diagnostics | 2 |
| Regulatory | 3 |
| Treasury and Finance | 1 |
| The National Advisory Group on Immunisation (NAGI) | 2 |
| Civil Society | 2 |
| National Department of Health | 6 |
| **Total** | **26** |
